# Gender Biases In Sports Concussions: A Differential Item Functioning Analysis into NCAA Female Athlete Symptom Presentation and Validity of the SCAT3 Checklist

**DOI:** 10.1101/2024.03.04.24303608

**Authors:** Rachel Edelstein, Sydney Cushing, Karen M. Schmidt, John Darrell Van Horn

## Abstract

**Background:** As a consequence of sports-related concussions, female athletes have been documented as reporting more symptoms than their male counterparts, in addition to incurring longer periods of recovery. However, the role of gender and its potential influence on symptom reporting and recovery outcomes in concussion management has not been completely explored.

**Study Design:** This study investigates potential differential item functioning (DIF) related to gender biases within the SCAT3 symptom severity checklist. The data was obtained from the Federal Interagency of Traumatic Brain Injury Research (FITBIR), which included information from the 2014-2017 NCAA and DoD CARE Consortium. A total of 1,258 NCAA athletes (*n*=473 females and *n*=785 males) SCAT3 Symptom Severity sub-scores were analyzed across five time points post-concussion: less than six hours post-injury, 24-48 hours post-injury, asymptomatic, unrestricted return to play, and at 6 months.

**Results:** During the recovery phase, women experienced more headaches, pressure in the head, and fatigue than male athletes. Overall, both male and female athletes had equivalent knowledge of concussions, and there was no significant difference in symptom-reporting ability for most items, including emotional-related symptoms. Only during the unrestricted return to play phase were group-level differences detected, with females being more likely to report more severe symptoms than males. However, upon further analysis, it was discovered females exhibit a relatively high difficulty level reporting symptom severity beyond ‘Mild’, therefore the group-level DIF may result from gender biases within the checklist.

**Conclusion:** The present analysis posits that Differential Item Functioning (DIF) in specific symptoms may lead to gender bias. The findings of this study reveal that female athletes tend to exhibit symptomatic behavior upon returning to play, a phenomenon consistent with prior research. However, the possible DIF may provoke biases due to unreliable reporting measures within the SCAT3 symptom severity checklist. Furthermore, explaining why recent literature reports that female athletes do not present as symptomatic upon return to play. Additional research is warranted to determine whether females genuinely experience more symptoms or whether the presence of these potential assessment gender biases obstructs the manifestation of asymptomatic recoveries.

## Introduction

Each year, over 460,000 student-athletes participate in collegiate sports competitions organized by the National Collegiate Athletic Association (NCAA)(Zuckerman et al., 2014). During the school years of 2009 to 2010 and 2013 to 2014, on average, 4.47 per 10,000 athletes experienced concussion exposures, which computes to 10,560 concussions annually, with women’s soccer being the second highest sport and rates in women’s soccer and volleyball having the most increased (Chandran et al., 2022). To manage and diagnose a sports-related concussion, the NCAA uses a unisex battery of assessments, one being a symptomatic presentation (Kroshus et al., 2021; Meehan et al., 2013; Dick 2009; McCrory et al., 2012; Harmon et al., 2013). However, several research studies have revealed that males and females exhibit different symptomatic patterns, both pre- and post-concussion, with females experiencing more reported concussions and prolonged recoveries. These inconsistent findings have led to the lack of clarity on the influence of gender on concussion management as well as recovery lengths (Broshek et al., 2005; Brown et al., 2015; Cantu & Register-Mihalik, 2011; Randolph et al., 2009).

If, as recent data suggest, a gender difference in sport-related concussion symptom reporting, it is still unclear why these differences arise. This is particularly consequential as symptom reporting plays a significant role in the decision-making process pertaining to return-to-play. A systematic review further examined gender differences in symptom reporting (D’Lauro et al., 2022), revealing that while it is unclear why women reported more symptoms, this possibly factored into why it took females a day longer to recover than male athletes. Similarly, one of the largest female-only analyses investigated this phenomenon within the NCAA and DoD CARE Consortium, revealing that asymptomatic presentation throughout recovery may not be a feasible expectation for female athletes, possibly due to limitations within current concussion management practices (Brown et al., 2015). Further investigation has indicated that clinical concussion assessments may be subject to sampling bias, as current assessments have been constructed around studies involving mostly male participants. (Gessel et al., 2007; Granito, 2002; Kieffer et al., 2021; Mihalik et al., 2009; O’Connor et al., 2017; Schatz et al., 2011; Wallace et al., 2017; Zuckerman et al., 2014). Therefore, generalizing head-injury and concussion assessment tools developed in males to the female athlete population is likely to present imprecise results.

Among the most widely used assessments is the Sport Concussion Assessment Tool (SCAT), (Echemendia et al., 2017) which is used to evaluate concussions in sideline, clinic, and hospital settings (McCrory et al., 2009, 2013; Pallant & Tennant, 2007). The SCAT combines several assessments, including a 22-item-graded symptom checklist, the Standard Assessment for Concussion (SAC), and a modified Balance Error Scoring System (mBESS) (Echemendia et al., 2017; Garcia et al., 2020). These components, initially chosen through consensus based on clinical experience and existing evidence, represent a robust combination of tests sensitive to concussion (Guskiewicz et al., 2013). Despite revisions to the SCAT, the symptom checklist has not been altered to include gender since the creation of the SCAT during the Second International Conference on Concussion in Sport in 2004, which is now in its sixth iteration (Yengo-Kahn et al. 2016; Chin et al., 2016).

Current concussion assessments and policies, to our knowledge, have not made any adjustments on how to manage gender in clinical settings. To address this issue, Item Response Theory (IRT) has emerged as a helpful approach, particularly with self-reported measures such as symptom reporting. IRT has recently gained popularity in abbreviating scales for concussion assessment and has been used to abbreviate various health-related and concussion outcomes while maintaining validity (Hamel et al., 2016; Heck et al., 2023; Langer et al., 2008). IRT can quantify gender-specific, shortened versions of multifaceted concussion assessments that are nearly as informative as longer ones (Angoff, 1981). In turn, identifying items that function differently across subgroups makes it a useful method for investigating gender differences in symptom reporting (Bollmann et al., 2018). IRT analysis is employed when a set of questionnaire items (or items from an administered scale) are intended to be summed together to provide a total score, which may include several subscale totals and an overall score, then be applied in the development of a new scale, where it is possible to design the item set to fit the model expectations from the outset.

Differential Item Functioning (DIF). DIF is particularly useful in identifying gender differences in symptom reporting (Kroshus et al., 2021; Langer et al., 2008; Tennant & Conaghan, 2007), as DIF can extract nuance gender differences in endorsing a given symptom after controlling for the overall scale score (Langer et al. 2008). The use of DIF has become an integral part of determining the validity and reliability of standardized tests, as DIF is not evident in symptoms when individuals from different groups have varying probabilities of responding in a certain way (Angoff, 1981; Langer et al., 2008; the EORTC Quality of Life Group and the Quality of Life Cross-Cultural Meta-Analysis Group et al., 2010). Rather, DIF becomes apparent when individuals from varying groups possessing the same true ability levels have varying probabilities of responding in a certain way. If, for example, in a symptom test, boys display higher probability of reporting symptoms more often than girls of equal ability level because the contents in the test items are biased against girls, then the assumption of this model includes unidimensionality and local independence (Ajeigbe & Afolabi, 2014). Unidimensionality occurs when each of the items in a test measures a single trait, for example, are all 22 symptoms on the SCAT3 symptom checklist measuring the latent trait severity recognition ability, which, in principle assumes local independence. Local independence is achieved when the probability to respond to items is independent of one another, suggesting that a response is based on the influence by no other symptom in the test.

The current study aims to investigate the SCAT3 symptom scores at various points during an athlete’s recovery process to identify any potential DIF at the item and group level that could be related to gender bias. Exploring potential DIF at the item and group level can reveal gender differences in symptom severity scoring that could impact the assessment tool’s validity. Ultimately, this study aims to provide insights into the potential impact of gender bias on SCAT3 Symptom Severity Checklist to advocate for the improvement of the accuracy and fairness of concussion assessments for female athletes.

## Methods

Data from the NCAA and DoD CARE Consortium was obtained from the Federal Interagency of Traumatic Brain Injury Research (FITBIR). From 2014-2017, 35,00 student-athletes and service cadets were from 26 different institutions, 4 US service academies, and over 15 different sports were collected. Within the CARE Consortium (CARE Consortium Investigators et al., 2017) approximately 13,009 military cadets and 21,006 student-athletes from NCAA Division 1-Division 3 completed baseline preseason testing, with 8,356 female athletes included (45.1%). All athletes met the following criteria: (i) they were identified with a diagnosed sports-related concussion, and (ii) data at all five desired time points. A total of five time points were examined: (i) less than six hours post-injury, (ii) 24-48 hours post-injury, (iii) asymptomatic, (iv) unrestricted return to play, and (v) at 6 months. Consequently, N=1,258 NCAA athletes met the inclusion criteria, with 473 female subjects (n= 473) and 785 male subjects (n=785). The specific sports included in this assessment are known for high incidences of concussion, including men’s football and men’s and women’s ice hockey, lacrosse, soccer, and rugby.

### Outcome Measures

The SCAT3 is a neurocognitive and symptom severity checklist based on a 7-point Likert scoring (*0-No symptoms, 1-2-Mild, 3-4-Moderate, 5-6-Severe)* (McCrory et al., 2009, 2013; Pallant & Tennant, 2007). For each item, a higher score indicates a more severe symptom (Echemendia et al., 2017). The SCAT3 was created to improve the original SCAT to assist in deciding when an athlete can safely return to play. The 22 items included were: *Headache*, *Pressure in the Head*, *Neck Pain*, *Nausea/Vomiting*, *Dizziness*, *Blurry Vision*, *Balance Problem*, *Sensitivity to Light*, *Sensitivity to Noise*, *Feeling Slowed Down*, *Feel in a Fog*, *Don’t Feel Right*, *Difficulty Concentrating*, *Difficulty Remembering*, *Fatigue/Low Energy*, *Confusion*, *Drowsiness*, *Trouble Falling Asleep*, *More Emotional*, *Irritable*, *Sadness*, and *Anxious*. The total score for each symptom is added to create a score out of 132, along with a score based on the total number of symptoms exhibited out of 22 (Begasse de Dhaem et al., 2017). Item response scales were rescaled to decompress the number of categories, as seen in Table 1, which reports assessment scoring scales, recoding measures, and corresponding adjusted response categories. The justification for compressing the response categories is to reduce categories with the same meaning despite different numbers (*e.g*., Mild = 1 and 2).

**Table 1:**
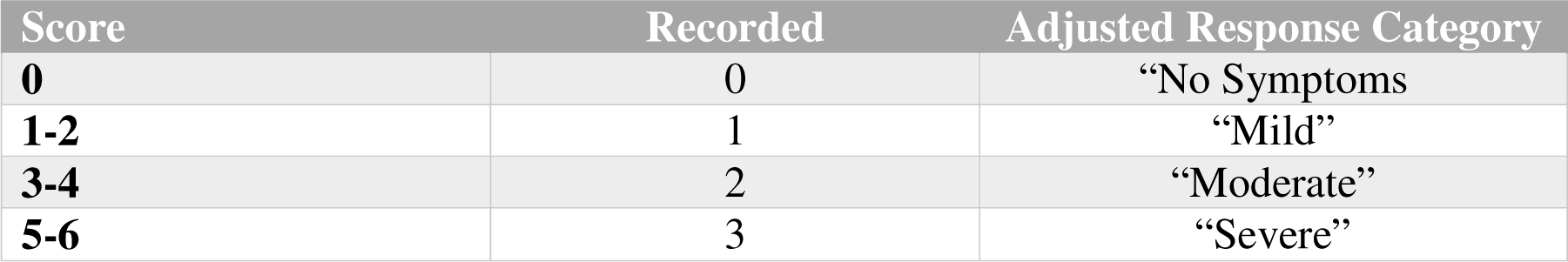
Assessment Adjust Response Categories for the Standardized Concussion Assessment Tool 3 (SCAT3)

### Statistical Analysis

This analysis used a model-based likelihood ratio test to identify DIF in a Partial Credit Model (PCM) through Winsteps Rasch Analysis and Methods Computer Software (Linacre, J. M. 2023). The PCM was chosen as it utilized flexible and well-suited Likert-type data such as the SCAT3 Symptom Severity Checklist. This item response modeling-based approach assumes the null hypothesis that the parameters for a particular item do not differ between groups. This method has an advantage in it isolates the parameters of an item, fitting a model with the parameters, allowing them to vary freely between groups, parameters constrained to be equal between groups, and uses as a test statistic that computes the difference between the loglikelihood values for the two models multiplied by −2 (Bollmann et al., 2018). The Winsteps Program anchors the item location, or difficulty (*b*) mean to zero. This compares the person’s ability or trait level scores, which are free to vary, in reference to the item scaling. Positive trait scores signify relatively higher performance, while negative trait scores indicate relatively lower performance, in reference to the items.

Item difficulty difference logits (*d)* was computed to see the impact on DIF within the SCAT3 symptom checklist and if it can be related to gender-bias. First, the widely used Mantel-Hazel (Dorans & Holland, 1992) method, which is a log-odds estimators of DIF size and significance from crosstabs of observations of the two genders. Second, DIF is estimated on a logit-difference, or logistic regression method, which estimates the difference between the symptom difficulties for the two groups, holding everything else constant. Post-hoc tests were done to prevent alpha inflation and Type I error brought about by multiple comparisons and to set the alpha level to *a*= 0.05, the Benjamini-Hochberg (B-H) procedure was utilized (Benjamini & Hochberg, 1995). The other comparison values are adjusted based on the rank order of the magnitudes for the observed p-values’ magnitudes, all falling within the range, based on the original statistical significance of DIF, a < 0.05.

DIF at the group level, or DGF, was computed for the interactions between classification groups of persons. The DGF Contrast will report the item’s difficulty difference between genders. DGF shows the difference in difficulty of the item between the two genders. The null hypothesis is that the two estimates are identical except for measurement error. Statistically significance DGF on an item, p ≤ 0.05. Therefore, a significant and positive DGF contrast indicates that the group-level differences which suggests more difficult for female athletes.

A Partial Credit Model (PCM; Masters, 1982 was used to estimate item category thresholds and corresponding expected posterior reliability at each time point. A PCM model estimates delta parameters (δ_ij_) are thresholds in which represent category difficulties and an item difficulty parameter (*b*) (Hays et al., 2000). Item difficulty (*b*) is computed by taking an average of all of the deltas (δ_ij_). As mentioned, an item reduction analysis was not performed, nor was it the focus of this analysis. The PCM was important to utilize as it demonstrated the current quality of the assessment without abbreviation; in turn, the PCM analysis will be used if DIF at the group level occurs. This will help determine whether each gender could accurately report symptom severity based on symptom difficulty. Posterior reliability measured by a Pearson correlation *(r*) coefficient (Akoglu, 2018), was also computed for both genders and all five time points.

Lastly, the high degree of unidimensionality was established before this analysis, which has demonstrated that the SCAT has a level of unidimensionality which implies that general concussion symptom severity is a coherent construct and could be estimated precisely using a subset of items from the full instrument. The present analysis assumes unidimensionality based on this validated bifactor approach (Nelson et al., 2018; Wilmoth et al., 2020; Brett et al., 2020). This approach is grounded on data indicating that concussion symptoms, as evaluated 24-48 hours post-injury by the SCAT3, are essentially unidimensional. A single underlying dimension, or a total symptom severity score, accounts for 96% of the reliable variance in ratings.

## Results

At the item level, statistically significant DIF (See Table 2) was detected solely during the initial three time-intervals. Specifically, Item difficulty difference *(d)* that impacted DIF within six hours, male athletes experienced greater difficulty in managing somatic symptoms, including Headache (*d*= −0.27, p < 0.001), pressure in the Head (*d*= −0.29, p = 0.04), and Sensitivity to Light (*d*= −0.36, p = 0.04). Two of these symptoms exhibited notable DIF. Within the 24-48 hour time frame, we continue to see the same clinically relevant trends as before, with males experiencing greater difficulty in recognizing the severity of Pressure in Head (*d*= 0.2, p = 0.04), and females seem to experience greater difficulty cognitive based symptoms identifying symptoms such as Don’t Feel Right (*d*= 0.29, *p* < 0.001). Additionally, More Emotional (*d*= −0.83, p <0.001) was significantly more difficult for males than females in the emotional domain. While the other symptoms in the emotional domain did not show significant DIF, they suggested difficulty for both genders. Finally, during the asymptomatic period, only one symptom showed DIF within the cognitive domain with feeling fog (0.41, p < 0.001), indicating greater difficulty for males. DIF was not detected past the asymptomatic period.

**Table 2:** Differential Item Functioning (DIF) for the SCAT3.

| Item | $d_{<6\text{hrs}}$ | $P\text{-value}$ | $a$ | $d_{24-48\text{hrs}}$ | $P\text{-value}$ | $a$ | $d_{\text{Asymptomatic}}$ | $P\text{-value}$ | $a$ | $d_{\text{Unrestricted}}$ | $P\text{-value}$ | $a$ | $d_{6\text{ months}}$ | $P\text{-value}$ | $a$ |
| --- | --- | --- | --- | --- | --- | --- | --- | --- | --- | --- | --- | --- | --- | --- | --- |
| Headache | **-0.27** | **<.001** | **<0.001\*\*** | -0.08 | 0.09 | 0.35 | 0.08 | 0.71 | 0.84 | 0.03 | 0.67 | 0.84 | -0.48 | 0.01 | 0.13 |
| Pressure in Head | -0.29 | 0.00 | 0.04* | -0.20 | 0.00 | 0.04 | -0.13 | 0.38 | 0.75 | 0.07 | 0.87 | 0.91 | -0.37 | 0.29 | 0.70 |
| Neck Pain | 0.19 | 0.59 | 0.84 | -0.11 | 0.13 | 0.42 | 0.18 | 0.75 | 0.85 | 0.63 | 0.23 | 0.61 | 0.39 | 0.33 | 0.71 |
| Nausea/Vomiting | -0.07 | 0.55 | 0.84 | -0.26 | 0.04 | 0.21 | 0.73 | 0.21 | 0.59 | -0.74 | 0.51 | 0.84 | -0.67 | 0.88 | 0.91 |
| Dizziness | 0.1 | 0.30 | 0.70 | 0.03 | 0.52 | 0.84 | 0.08 | 0.71 | 0.84 | 1.20 | 0.23 | 0.61 | -1.75 | 0.07 | 0.31 |
| Blurry Vision | 0.01 | 0.63 | 0.84 | 0.30 | 0.02 | 0.13 | -0.47 | 0.48 | 0.84 | 0.48 | 0.57 | 0.84 | 0.28 | 0.38 | 0.74 |
| Balance Problem | 0.26 | 0.03 | 0.17 | 0.19 | 0.10 | 0.38 | -0.28 | 0.70 | 0.84 | -0.95 | 0.59 | 0.84 | 1.47 | 0.03 | 0.20 |
| Sensitivity Light | **-0.36** | **0.00** | **0.04\*** | 0.04 | 0.60 | 0.84 | -0.32 | 0.49 | 0.84 | 0.14 | 0.83 | 0.90 | 0.46 | 0.43 | 0.78 |
| Sensitivity Noise | -0.24 | 0.09 | 0.35 | -0.07 | 0.68 | 0.84 | 0.04 | 0.39 | 0.75 | 0.17 | 0.66 | 0.84 | 0.34 | 0.86 | 0.91 |
| Feel Slow Down | 0.1 | 0.13 | 0.42 | 0.02 | 0.54 | 0.84 | -0.04 | 0.73 | 0.84 | 0.31 | 0.22 | 0.61 | -0.35 | 0.88 | 0.91 |
| Feel Fog | 0.24 | 0.54 | 0.84 | 0.29 | 0.02 | 0.13 | **0.41** | **< 0.001** | **0.00** | 0.10 | 0.54 | 0.84 | -0.05 | 0.79 | 0.88 |
| Don't Feel Right | 0.13 | 0.05 | 0.27 | **0.29** | **<.001** | **<0.001\*\*** | 0.12 | 0.07 | 0.31 | 0.85 | 0.16 | 0.50 | 0.35 | 0.13 | 0.42 |
| Difficulty Concentrating | -0.04 | 0.70 | 0.84 | 0.09 | 0.13 | 0.42 | -0.58 | 0.03 | 0.20 | 0.06 | 0.65 | 0.84 | -0.09 | 0.29 | 0.70 |
| Difficulty Remembering | **0.57** | **< 0.001** | **< 0.001\*\*** | 0.26 | 0.00 | 0.05* | -0.01 | 0.72 | 0.84 | -0.07 | 0.49 | 0.84 | -0.28 | 0.78 | 0.88 |
| Low Energy | -0.32 | 0.04 | 0.20 | -0.15 | 0.07 | 0.31 | 0.14 | 0.52 | 0.84 | 0.05 | 0.67 | 0.84 | 0.03 | 0.89 | 0.91 |
| Confusion | **0.76** | **<.001** | **<0.001\*\*** | 0.22 | 0.01 | 0.08 | -0.16 | 1.00 | 1.00 | -2.17 | 0.32 | 0.71 | 1.60 | 0.34 | 0.73 |
| Drowsiness | -0.18 | 0.19 | 0.57 | -0.10 | 0.52 | 0.84 | 0.31 | 0.36 | 0.73 | -0.39 | 0.36 | 0.73 | 0.44 | 0.28 | 0.70 |
| Trouble Falling Asleep | -0.09 | 0.68 | 0.84 | 0.32 | 0.14 | 0.44 | 0.24 | 0.71 | 0.84 | -0.12 | 0.97 | 0.98 | 0.22 | 0.41 | 0.77 |
| More Emotional | -0.42 | 0.07 | 0.31 | **-0.83** | **<.001** | **<0.001\*\*** | -0.66 | 0.11 | 0.40 | -2.15 | 0.02 | 0.15 | -0.48 | 0.32 | 0.71 |
| Irritable | 0.1 | 0.55 | 0.84 | -0.06 | 0.62 | 0.84 | -0.17 | 0.33 | 0.71 | -0.51 | 0.86 | 0.91 | -0.03 | 0.71 | 0.84 |
| Sadness | -0.21 | 0.23 | 0.61 | -0.44 | 0.01 | 0.13 | 0.04 | 0.80 | 0.89 | 0.05 | 0.58 | 0.84 | -0.43 | 0.61 | 0.84 |
| Nervous/Anxious | -0.09 | 0.30 | 0.70 | -0.03 | 0.65 | 0.84 | -0.02 | 0.74 | 0.85 | -0.65 | 0.10 | 0.38 | -0.41 | 0.35 | 0.73 |

During unrestricted return to play, the individual variability in symptom difficulty did not significantly impact the differential item functioning (DIF). However, the total symptom severity scores demonstrated that symptom difficulty varied significantly between genders (Table 3). Females reported symptom severity scores that were, on average, 38% higher than males (*d* = −0.38, p < 0.001). The presence of this DIF signifies that certain symptoms might be measuring different traits among genders, which could potentially lead to biased outcomes and misguided symptom severity scores.

**Table 3:** Differential Group Function Interaction Bias.

| TIME PERIOD | DGF<br>CONTRAST | JOINT<br>S.E. | Rasch-Welch<br>T-Value | P-value |
| --- | --- | --- | --- | --- |
| < 6 hours | 0.00 | 0.03 | 0.00 | 1.00 |
| 24-48 hours | 0.00 | 0.02 | 0.00 | 1.000 |
| Asymptomatic | 0.08 | 0.05 | -1.55 | 0.1207 |
| Unrestricted Return to Play | **-0.38** | **0.10** | **-3.85** | **0.0001** |
| 6 Months | -0.13 | 0.07 | -1.75 | 0.0794 |

As seen in Table 4, the assessment’s reliability is at its lowest during the unrestricted return to play (*r*=0.47 vs *r*=0.31) but at its highest during the 24-48 for both females and males (*r*=0.92 vs *r*=0.92). In Table 5, to further investigate if the DIF detected at the group level can be related to gender, trait levels were an unrestricted return to play. The Item(s) contained zero observations from males and were therefore dropped: these were Nausea/Vomiting, Balance Problem, Confusion, More Emotional, and Irritable. Table 5 displays the item location parameters and category thresholds for the PCM model during the unrestricted return to play period; The analysis revealed that females item location (b) is equal to or less than the difficulty of reporting mild symptom (δ1), the ability to report symptoms beyond this severity level would be difficult.

**Table 4:** Posterior Pearson Reliability (r)

| Sex of Athlete | < 6 hours | 24-48 hours | Asymptomatic | Unrestricted | 6 Months |
| --- | --- | --- | --- | --- | --- |
| **Females** | 0.90 | 0.92 | 0.65 | 0.47 | 0.6 |
| **Males** | 0.92 | 0.92 | 0.59 | 0.31 | 0.50 |

**Table 5:** Unrestricted Return to Play Item Category Difficulty Thresholds.

| <u>Males</u> | Item<br>difficulty<br>(*b*) | Mild<br>(□<sub>1</sub>) | Moderate<br>(□<sub>2</sub>) | <u>Females</u> | Item<br>difficulty<br>(*b*) | Mild<br>(□<sub>1</sub>) | Moderate<br>(□<sub>2</sub>) |
| --- | --- | --- | --- | --- | --- | --- | --- |
| Headache | -1.86 | -1.86 | - | Headache | -0.48 | -2.08 | 1.12 |
| Pressure in Head | -1.37 | -1.37 | - | Pressure in Head | -0.23 | -1.15 | 0.68 |
| Neck Pain | -1.63 | -1.63 | - | Neck Pain | -1.09 | -1.09 | - |
| Dizziness | 0.92 | 0.92 | - | Nausea/Vomiting | 1.78 | 1.78 | - |
| Blurry Vision | 1.72 | 1.72 | - | Dizziness | 1.04 | 1.04 | - |
| Sensitivity Light | -0.24 | -0.24 | - | Blurry Vision | 1.04 | 1.04 | - |
| Sensitivity Noise | 0.92 | 0.92 | - | Balance Problem | 1.04 | 1.04 | - |
| Feel Slow Down | -0.24 | -0.24 | - | Sensitivity Light | -0.20 | -0.20 | - |
| Feel Fog | 0.06 | 0.06 | - | Sensitivity Noise | 1.04 | 1.04 | - |
| Don't Feel Right | 0.06 | 0.06 | - | Feel Slow Down | 0.60 | 0.60 | - |
| Difficult<br>Concentrating | 0.16 | -0.10 | 0.43 | Feel Fog | 0.60 | 0.60 | - |
| Difficult<br>Remembering | 0.06 | 0.06 | - | Don't Feel Right | 0.60 | 0.60 | - |
| Low Energy | -1.07 | -1.07 | - | Difficulty<br>Concentrating | -0.39 | -0.39 | - |
| Drowsiness | -0.71 | -0.71 | - | Difficulty<br>Remembering | 0.06 | 0.97 | -0.84 |
| Trouble Falling<br>Asleep | 0.06 | 0.06 | - | Low Energy | -0.62 | -1.08 | -0.16 |
| Sadness | 1.72 | 1.72 | - | Confusion | 1.04 | 1.04 | - |
| Nervous/Anxious | 0.33 | 0.73 | -0.06 | Drowsiness | -0.15 | -0.77 | 0.46 |
|  |  |  |  | Trouble Falling<br>Asleep | -0.20 | -0.20 | - |
|  |  |  |  | More Emotional | -0.07 | -0.26 | 0.13 |
|  |  |  |  | Irritable | -0.20 | -0.20 | - |
|  |  |  |  | Sadness | 0.60 | 0.60 | - |
|  |  |  |  | Nervous/Anxious | 0.02 | 0.02 | - |

Figure 1 visually displays the outcome of the PCM analysis through a Person-Item Map. These plots show each genders abilities and item difficulties respectively along the same latent dimension. Dark circles can be that of the item difficulty and the open circles are category difficulty. For females a total of 6 symptoms, Headache, Neck Pain, Difficulty Remembering, Low Energy, Drowsiness and More Emotional. were more easily distinguishable between categories, however, the remaining 16 symptoms were too difficult for females to accurate report symptom severity. Additionally, 12 out of the 22 symptoms item difficulty was b < 0, indicating difficultly to categorize past ‘Mild’. For male athletes, the item difficulties for Headache, Pressure in Heath, Neck Pain, Sensitivity to light, Feeling Slowed Down, Difficulty Remembering, and Low Energy were relatively easy to identify and contained zero observations in the Moderate or Severe Category.

**Figure 1:**
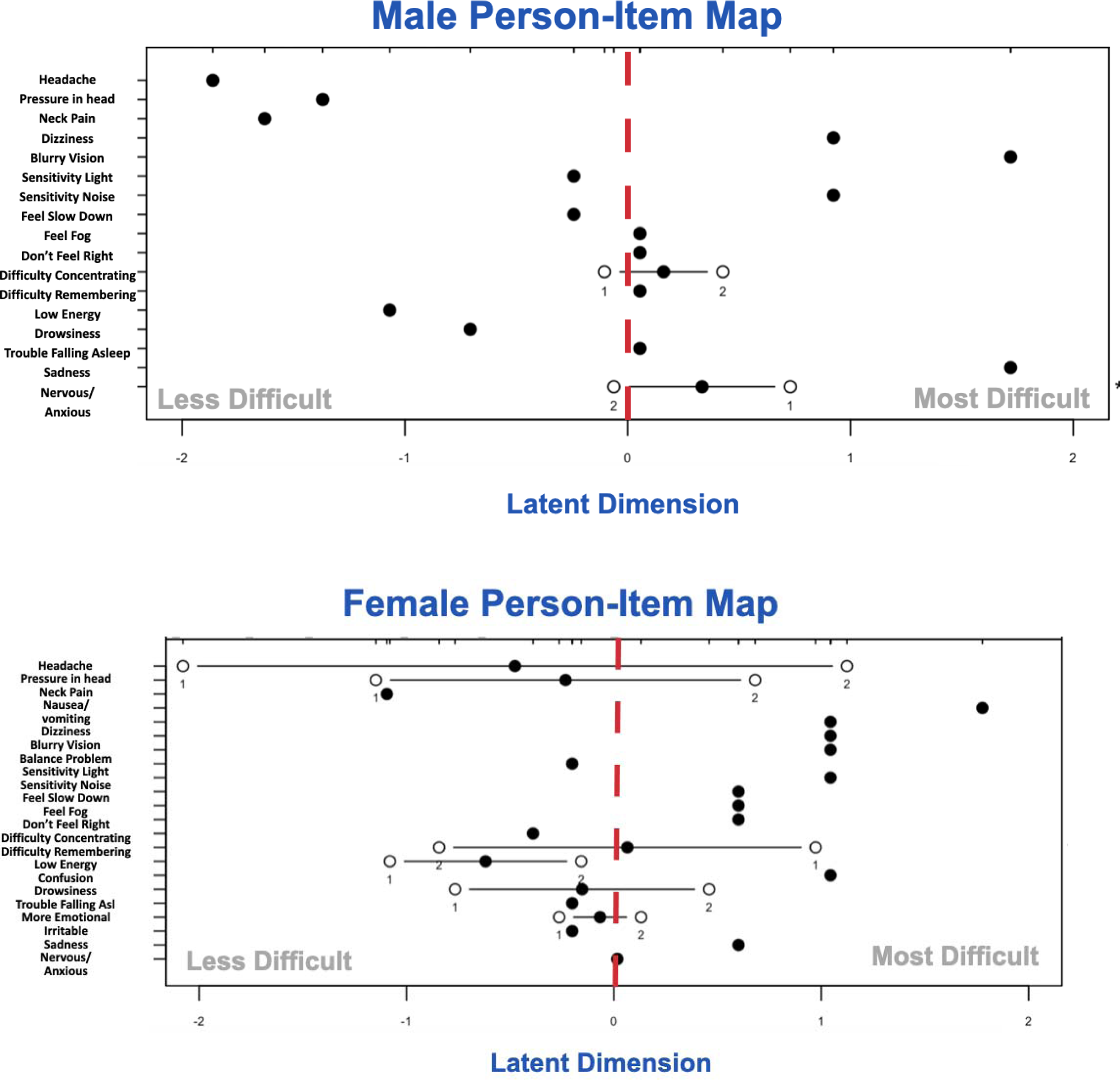
Person-Item Mappings. The person-item mappings for male (top) and for female (bottom) athletes, which display the location of person abilities and item difficulties respectively along the same latent dimension. Locations of item difficulties are displayed with solid circles and thresholds (1= Mild, 2=Moderate) of adjacent category locations with open circles. Dark circles which are greater than zero are considered more difficult symptoms.

## Discussion

Based on the findings of this IRT analysis, somatic and cognitive concussion symptoms appear to be more easily identifiable within 48 hours of the injury, regardless of gender, apart from symptoms of nausea and/or vomiting. Prior studies using the IRT model indicate that a condensed variant of a symptom checklist consisting of 3, 9, or 12-item checklists is just as effective as a longer version containing 22 items in identifying concussion (Randolph et al., 2009; Wilmoth et al., 2020). These studies found similar cognitive and somatic symptoms, such as feeling slow, foggy, and not right, being most identifiable. This is consistent with the present work, finding the most difficult symptoms for females to identify somatic symptoms (Headache, Pressure in Head, Neck pain, Nausea/Vomiting, Dizziness, Balance problems, Sensitivity to light, Sensitivity to noise, Fatigue or low energy) whereas emotional symptoms (Feeling like “in a fog”, Difficulty Concentrating, Difficulty Remembering, Confusion) manifested the most difficult domain. Additionally, in line with these studies, Headache was the best-performing item for low-severity symptoms, which supports this item’s important role in concussion assessment. This analysis further confirms that SCAT symptom checklist has a high degree of coherence, suggesting that general concussion symptom severity can be accurately estimated using a subset of items from the somatic and cognitive domain of the full instrument (Brett et al., 2020; Wilmoth et al., 2020).

Despite limited items detecting DIF, also aligning with these studies, this analysis revealed that emotion-related symptoms were not easily identified in either gender, particularly over 48 hours (Chin et al., 2016; Covassin & Elbin, 2011; Granito, 2002; Preiss-Farzanegan et al., 2009). Asking an athlete to report their emotional symptoms (More Emotional, Irritability, Sadness, Nervous or Anxious) concerning their concussion may be too non-specific in relation to concussion in both genders; thus, symptoms such as “Are you Sad” or “Feeling Irritable” may be too vague or difficult to report accurately. Given the cofounding of other variables within an athlete’s life (*e.g.*, academic pressures, family, relationships, and so on), this may be a reason why these symptoms are more difficult to identify; therefore, removing the emotional domain could enhance the accuracy of symptom reporting and improve recovery and rehabilitation outcomes.

Results indicated that there may be a disparity in how male and females athletes report their symptoms during the final phase before returning to play. Which can arguably one of the most important time frames before an athlete returns to play. This disparity could be a contributing factor to the recent studies indicating that female athletes do not report symptoms as frequently as their male counterparts during this phase. The analysis also uncovered challenges with accurate symptom reporting for both male and female athletes during this phase. Additionally, in line with previous findings (Nelson et al., 2018; Wilmoth et al., 2020), it was discovered that the SCAT3 symptom checklist may not be a reliable assessment measure when applied to female athletes and their returning to play as the reliability of the assessment was found exceptionally unreliable. The data also revealed that male athletes did not perceive five symptoms at all, suggesting that they either did not experience these symptoms or were underreporting them. By considering these discrepancies and assessment issues, it is possible that gender biases may be influencing the results and creating an inaccurate perception of female athletes appearing more symptomatic during the return to play phase.

In determining clinical symptom presentation post-concussion, it will be important to consider how psychological and social factors influence gender biases at different stages of recovery (Beran & Scafide, 2022; Caccese, et al. 2023; CARE Consortium Investigators et al., 2018; Lempke, Oldham, et al. 2023; Sinnott et al., 2023). Researchers may want to expand on the current Rasch Partial Credit Model to account for additional parameters to understand better how much external factors influence accurate symptom reporting. Rasch modeling will be a useful tool for researchers in the concussion field to evaluate the relationship between sociological pressures, such as reporting intentions, and diagnostic measures, such as symptom presentation on the variability of recovery length.

Futhermore, unlike basic regression modeling, this modeling approach can be from two different assessments (*e.g.*, ImPACT vs SCAT) and consider scale equating techniques (Kolen & Brennan, 2004; Simons et. al., 2022; Choi et. al., 2014). Rasch item response modeling approaches calibrate different scales and assessment onto the latent variable, thus accounting for measurement error and reducing potential bias (Kolen & Brennan, 2004). Previous studies attempted to validate this approach by in calibrating the SCAT and Rivermead Post Concussion Symptoms Questionnaire (RPQ), using fixed-parameter calibration allows for a single item calibration for all items, offering a more rigorously established cross-walk between the assessments. Findings revealed benefits of this modeling approach revealed more reliability and precision than other methods, which linked RPQ and SCAT total scores based on their relationship to the latent dimension (TBI-related symptom burden) that drives scores on both measures. Moreover, Rasch-based item response modeling is a robust, yet minimally used, modeling approach which can be beneficial in capturing this very important construct on the same scale as the items used to measure it. Until these assessments are analyzed to better reflect potential differential relationships of gender, it will be important to factor in certain symptomatic presentations when making return-to-play decisions, particularly for females.

## Study Limitations

This research was conducted on a specific group of athletes participating in contact sports in the NCAA, such as football, lacrosse, field/ice hockey, soccer, and rugby. It is important to note these results cannot be generalized to youth, high school, or professional sports. Therefore, it would be beneficial to conduct further studies on larger, national samples that include different age groups, contact and non-contact, as well as youth, collegiate, and professional sports to better to understand the psychometric properties of SCAT short forms as they pertain to female athletes.

## Conclusion

As a result of this IRT analysis, based on *N*=1,258 NCAA athletes, it is worth considering dthat the assumption that females experience more concussion symptoms and are at a higher risk of prolonged symptoms may not be entirely accurate. The findings of this analysis suggest that current concussion assessments may contain gender biases resulting in differences in symptom presentation, further justifying the idea that these concussion assessments based on male-based normative data cannot be generalized to the female athlete population. Moreover, these differences in symptom presentation may be causing female athletes to be considered as “Not Recovered” - potentially hindering their returning to field of play. Therefore, an investigation into constructing tailored, female-specific, symptom assessments may be an important first step in more accurately capturing the concussion recovery process in female athletes.

## Data Availability

All data produced in the present study are available upon reasonable request to the authors.

## References

Ajeigbe, T. O., & Afolabi, E. R. I. (2014). Assessing Unidimensionality and Differential Item Functioning in Qualifying Examination for Senior Secondary School Students, Osun State, Nigeria. World Journal of Education, 4(4), p30. 10.5430/wje.v4n4p30

Akoglu, H. (2018). User’s guide to correlation coefficients. Turkish Journal of Emergency Medicine, 18(3), 91–93. 10.1016/j.tjem.2018.08.001

Angoff, W. (1981). Use of Difficulty and Discrimination Indices for Detecting Item Bias DIF. Handbook of Methods for Detecting Test Bias., p96–116.

Begasse de Dhaem, O., Barr, W. B., Balcer, L. J., Galetta, S. L., & Minen, M. T. (2017). Post-traumatic headache: The use of the sport concussion assessment tool (SCAT-3) as a predictor of post-concussion recovery. The Journal of Headache and Pain, 18(1), 60. 10.1186/s10194-017-0767-5

Benjamini, Y., & Hochberg, Y. (1995). Controlling the False Discovery Rate: A Practical and Powerful Approach to Multiple Testing. Journal of the Royal Statistical Society: Series B (Methodological), 57(1), 289–300. 10.1111/j.2517-6161.1995.tb02031.x

Beran, K. M., & Scafide, K. N. (2022). Factors Related to Concussion Knowledge, Attitudes, and Reporting Behaviors in US High School Athletes: A Systematic Review. Journal of School Health, 92(4), 406–417. 10.1111/josh.13140

Bollmann, S., Berger, M., & Tutz, G. (2018). Item-Focused Trees for the Detection of Differential Item Functioning in Partial Credit Models. Educational and Psychological Measurement, 78(5), 781–804. 10.1177/0013164417722179

Brett, B. L., Kramer, M. D., McCrea, M. A., Broglio, S. P., McAllister, T. W., Nelson, L. D., the CARE Consortium Investigators, Hazzard, J. B., Kelly, L. A., Ortega, J., Port, N., Pasquina, P. F., Jackson, J., Cameron, K. L., Houston, M. N., Goldman, J. T., Giza, C., Buckley, T., Clugston, J. R., … Susmarski, A. (2020). Bifactor Model of the Sport Concussion Assessment Tool Symptom Checklist: Replication and Invariance Across Time in the CARE Consortium Sample. The American Journal of Sports Medicine, 48(11), 2783–2795. 10.1177/0363546520946056

Broshek, D. K., Kaushik, T., Freeman, J. R., Erlanger, D., Webbe, F., & Barth, J. T. (2005). Sex differences in outcome following sports-related concussion. Journal of Neurosurgery, 102(5), 856–863. 10.3171/jns.2005.102.5.0856

Brown, D. A., Elsass, J. A., Miller, A. J., Reed, L. E., & Reneker, J. C. (2015a). Differences in Symptom Reporting Between Males and Females at Baseline and After a Sports-Related Concussion: A Systematic Review and Meta-Analysis. Sports Medicine, 45(7), 1027–1040. 10.1007/s40279-015-0335-6

Cantu, R. C., & Register-Mihalik, J. K. (2011). Considerations for Return-to-Play and Retirement Decisions After Concussion. PM&R, 3(10S2). 10.1016/j.pmrj.2011.07.013

CARE Consortium Investigators, Broglio, S. P., Katz, B. P., Zhao, S., McCrea, M., & McAllister, T. (2018). Test-Retest Reliability and Interpretation of Common Concussion Assessment Tools: Findings from the NCAA-DoD CARE Consortium. Sports Medicine, 48(5), 1255–1268. 10.1007/s40279-017-0813-0

Chandran, A., Boltz, A. J., Morris, S. N., Robison, H. J., Nedimyer, A. K., Collins, C. L., & Register-Mihalik, J. K. (2022). Epidemiology of Concussions in National Collegiate Athletic Association (NCAA) Sports: 2014/15-2018/19. The American Journal of Sports Medicine, 50(2), 526–536. 10.1177/03635465211060340

Chiang Colvin, A., Mullen, J., Lovell, M. R., Vereeke West, R., Collins, M. W., & Groh, M. (2009). The Role of Concussion History and Gender in Recovery from Soccer-Related Concussion. The American Journal of Sports Medicine, 37(9), 1699–1704. 10.1177/0363546509332497

Chin, E. Y., Nelson, L. D., Barr, W. B., McCrory, P., & McCrea, M. A. (2016). Reliability and Validity of the Sport Concussion Assessment Tool–3 (SCAT3) in High School and Collegiate Athletes. The American Journal of Sports Medicine, 44(9), 2276–2285. 10.1177/0363546516648141

Choi, S. W., Schalet, B., Cook, K. F., & Cella, D. (2014). Establishing a common metric for depressive symptoms: linking the BDI-II, CES-D, and PHQ-9 to PROMIS depression. Psychological assessment, 26(2), 513–527. 10.1037/a0035768

Covassin, T., & Elbin, R. J. (2011). The Female Athlete: The Role of Gender in the Assessment and Management of Sport-Related Concussion. Clinics in Sports Medicine, 30(1), 125–131. 10.1016/j.csm.2010.08.001

Covassin, T., Elbin, R., Kontos, A., & Larson, E. (2010). Investigating baseline neurocognitive performance between male and female athletes with a history of multiple concussion. Journal of Neurology, Neurosurgery & Psychiatry, 81(6), 597–601. 10.1136/jnnp.2009.193797

Covassin, T., Swanik, C. B., Sachs, M., Kendrick, Z., Schatz, P., Zillmer, E., Kaminaris, C., Iverson, G., & Stearne, D. J. (2006). Sex differences in baseline neuropsychological function and concussion symptoms of collegiate athletes * Commentary * Commentary. British Journal of Sports Medicine, 40(11), 923–927. 10.1136/bjsm.2006.029496

Covassin, T., Swanik, C. B., & Sachs, M. L. (2003). Sex Differences and the Incidence of Concussions Among Collegiate Athletes. Journal of Athletic Training, 38(3), 238–244.

Davis-Hayes, C., Gossett, J. D., Levine, W. N., Shams, T., Harada, J., Mitnick, J., & Noble, J. (2017). Sex-specific Outcomes and Predictors of Concussion Recovery. Journal of the American Academy of Orthopaedic Surgeons, 25(12), 818–828. 10.5435/JAAOS-D-17-00276

Dick, R. W. (2009a). Is there a gender difference in concussion incidence and outcomes? British Journal of Sports Medicine, 43(Suppl_1), i46–i50. 10.1136/bjsm.2009.058172

D’Lauro, C., Jones, E. R., Swope, L. M., Anderson, M. N., Broglio, S., & Schmidt, J. D. (2022). Under-representation of female athletes in research informing influential concussion consensus and position statements: An evidence review and synthesis. British Journal of Sports Medicine, 56(17), 981–987. 10.1136/bjsports-2021-105045

Dorans, N. J., & Holland, P. W. (1992). DIF DETECTION AND DESCRIPTION: MANTEL-HAENSZEL AND STANDARDIZATION1,2. ETS Research Report Series, 1992(1). 10.1002/j.2333-8504.1992.tb01440.x

Echemendia, R. J., Meeuwisse, W., McCrory, P., Davis, G. A., Putukian, M., Leddy, J., Makdissi, M., Sullivan, S. J., Broglio, S. P., Raftery, M., Schneider, K., Kissick, J., McCrea, M., Dvorak, J., Sills, A. K., Aubry, M., Engebretsen, L., Loosemore, M., Fuller, G., … Herring, S. (2017). The Sport Concussion Assessment Tool 5th Edition (SCAT5). British Journal of Sports Medicine, bjsports-2017–097506. 10.1136/bjsports-2017-097506

Garcia, G.-G. P., Yang, J., Lavieri, M. S., McAllister, T. W., McCrea, M. A., Broglio, S. P., & on behalf of the CARE Consortium Investigators. (2020). Optimizing Components of the Sport Concussion Assessment Tool for Acute Concussion Assessment. Neurosurgery, 87(5), 971–981. 10.1093/neuros/nyaa150

Gessel, L. M., Fields, S. K., Collins, C. L., Dick, R. W., & Comstock, R. D. (2007). Concussions among United States high school and collegiate athletes. Journal of Athletic Training, 42(4), 495–503.

Granito, V. J. (2002). Psychological response to athletic injury: Gender differences. Journal of Sport Behavior, 25(3), 243+. Gale Academic OneFile.

Guskiewicz, K. M., Register-Mihalik, J., McCrory, P., McCrea, M., Johnston, K., Makdissi, M., Dvořák, J., Davis, G., & Meeuwisse, W. (2013). Evidence-based approach to revising the SCAT2: Introducing the SCAT3: Table 1. British Journal of Sports Medicine, 47(5), 289–293. 10.1136/bjsports-2013-092225

Hamel, J.-F., Sébille, V., Challet-Bouju, G., & Hardouin, J.-B. (2016). Partial Credit Model: Estimations and Tests of Fit with Pcmodel. The Stata Journal: Promoting Communications on Statistics and Stata, 16(2), 464–481. 10.1177/1536867X1601600212

Harmon, K. G., Drezner, J. A., Gammons, M., Guskiewicz, K. M., Halstead, M., Herring, S. A., Kutcher, J. S., Pana, A., Putukian, M., & Roberts, W. O. (2013). American Medical Society for Sports Medicine position statement: Concussion in sport. British Journal of Sports Medicine, 47(1), 15–26. 10.1136/bjsports-2012-091941

Hays, R. D., Morales, L. S., & Reise, S. P. (2000). Item Response Theory and Health Outcomes Measurement in the 21st Century: Medical Care, 38, II-28–II-42. 10.1097/00005650-200009002-00007

Heck, S. J., Acord-Vira, A., & Davis, D. R. (2023). Sex differences in college students’ knowledge of concussion and concussion education sources. Concussion, 8(3), CNC108. 10.2217/cnc-2023-0001

Kieffer, E. E., Brolinson, P. G., Maerlender, A. E., Smith, E. P., & Rowson, S. (2021). In-Season Concussion Symptom Reporting in Male and Female Collegiate Rugby Athletes. Neurotrauma Reports, 2(1), 503–511. 10.1089/neur.2021.0050

Kochick, V., Sinnott, A. M., Eagle, S. R., Bricker, I. R., Collins, M. W., Mucha, A., Connaboy, C., & Kontos, A. P. (2022). The Dynamic Exertion Test for Sport-Related Concussion: A Comparison of Athletes at Return-to-Play and Healthy Controls. International Journal of Sports Physiology and Performance, 17(6), 834–843. 10.1123/ijspp.2021-0258

Kolen, M. J., & Brennan, R. L. (2004). Test equating, scaling, and linking: Methods and practices. Springer. Retrieved from https://books.google.com/books?id=tusOwjb7LWsC

Kroshus, E., Lowry, S. J., Garrett, K., Hays, R., Hunt, T., & Chrisman, S. P. D. (2021a). Development of a scale to measure expected concussion reporting behavior. Injury Epidemiology, 8(1), 70. 10.1186/s40621-021-00364-4

Langer, M. M., Hill, C. D., Thissen, D., Burwinkle, T. M., Varni, J. W., & DeWalt, D. A. (2008a). Item response theory detected differential item functioning between healthy and ill children in quality-of-life measures. Journal of Clinical Epidemiology, 61(3), 268–276. 10.1016/j.jclinepi.2007.05.002

Langlois, J. A., Rutland-Brown, W., & Wald, M. M. (2006). The Epidemiology and Impact of Traumatic Brain Injury: A Brief Overview. Journal of Head Trauma Rehabilitation, 21(5), 375–378. 10.1097/00001199-200609000-00001

Linacre, J. (1965). WINSTEPS®. https://www.winsteps.com/index.htm

Lempke, L. B., Caccese, J. B., Syrydiuk, R. A., Buckley, T. A., Chrisman, S. P. D., Clugston, J. R., Eckner, J. T., Ermer, E., Esopenko, C., Jain, D., Kelly, L. A., Memmini, A. K., Mozel, A. E., Putukian, M., Susmarski, A., Pasquina, P. F., McCrea, M. A., McAllister, T. W., Broglio, S. P., … CARE Consortium Investigators. (2023). Female Collegiate Athletes’ Concussion Characteristics and Recovery Patterns: A Report from the NCAA-DoD CARE Consortium. Annals of Biomedical Engineering. 10.1007/s10439-023-03367-y

Lempke, L. B., Johnson, R. S., Schmidt, J. D., & Lynall, R. C. (2020). Clinical versus Functional Reaction Time: Implications for Postconcussion Management. Medicine & Science in Sports & Exercise, 52(8), 1650–1657. 10.1249/MSS.0000000000002300

Lempke, L. B., Oldham, J. R., Passalugo, S., Willwerth, S. B., Berkstresser, B., Wang, F., Howell, D. R., & Meehan, W. P. (2023). Influential Factors and Preliminary Reference Data for a Clinically Feasible, Functional Reaction Time Assessment: The Standardized Assessment of Reaction Time. Journal of Athletic Training, 58(2), 112–119. 10.4085/1062-6050-0073.22

Lempke, L. B., Passalugo, S., Baranker, B. T., Hunt, D., Berkstresser, B., Wang, F., Meehan, W. P., & Howell, D. R. (2022). Relationship and Latent Factors Between Clinical Concussion Assessments and the Functional Standardized Assessment of Reaction Time (StART). Clinical Journal of Sport Medicine, 32(6), e591–e597. 10.1097/JSM.0000000000001061

Lempke, L. B., Shumski, E. J., Prato, T. A., & Lynall, R. C. (2023). Reliability and Minimal Detectable Change of the Standardized Assessment of Reaction Time. Journal of Athletic Training, 58(6), 579–587. 10.4085/1062-6050-0391.22

Lidvall, H. F., Linderoth, B., & Norlin, B. (1974). Causes of the post-concussional syndrome. Acta Neurologica Scandinavica. Supplementum, 56, 3–144.

Masters, G.N. (1982). A Rasch model for partial credit scoring. Psychometrika, 47, 149–174.

McCrory, P., Meeuwisse, W., Aubry, M., Cantu, B., Dvořák, J., Echemendia, R., Engebretsen, L., Johnston, K., Kutcher, J., Raftery, M., Sills, A., Benson, B., Davis, G., Ellenbogen, R., Guskiewicz, K., Herring, S. A., Iverson, G., Jordan, B., Kissick, J., … Turner, M. (2013a). Consensus statement on Concussion in Sport – The 4th International Conference on Concussion in Sport held in Zurich, November 2012. Physical Therapy in Sport, 14(2), e1–e13. 10.1016/j.ptsp.2013.03.002

McCrory, P., Meeuwisse, W., Johnston, K., Dvorak, J., Aubry, M., Molloy, M., & Cantu, R. (2009). Consensus Statement on Concussion in Sport 3rd International Conference on Concussion in Sport Held in Zurich, November 2008. Clinical Journal of Sport Medicine, 19(3), 185–200. 10.1097/JSM.0b013e3181a501db

Meehan, W. P., Mannix, R. C., O’Brien, M. J., & Collins, M. W. (2013). The Prevalence of Undiagnosed Concussions in Athletes. Clinical Journal of Sport Medicine, 23(5), 339–342. 10.1097/JSM.0b013e318291d3b3

Mihalik, J. P., Ondrak, K. S., Guskiewicz, K. M., & McMurray, R. G. (2009). The effects of menstrual cycle phase on clinical measures of concussion in healthy college-aged females. Journal of Science and Medicine in Sport, 12(3), 383–387. 10.1016/j.jsams.2008.05.003

National Research Council (U.S.), Graham, R., Rivara, F. P., Ford, M. A., Spicer, C. M., & Institute of Medicine (U.S.) (Eds.). (2014). Sports-related concussions in youth: Improving the science, changing the culture. The National Academies Press.

Nelson, L. D., Kramer, M. D., Patrick, C. J., & McCrea, M. A. (2018). Modeling the Structure of Acute Sport-Related Concussion Symptoms: A Bifactor Approach. Journal of the International Neuropsychological Society, 24(8), 793–804. 10.1017/S1355617718000462

O’Connor, K. L., Baker, M. M., Dalton, S. L., Dompier, T. P., Broglio, S. P., & Kerr, Z. Y. (2017). Epidemiology of Sport-Related Concussions in High School Athletes: National Athletic Treatment, Injury and Outcomes Network (NATION), 2011–2012 Through 2013–2014. Journal of Athletic Training, 52(3), 175–185. 10.4085/1062-6050-52.1.15

Pallant, J. F., & Tennant, A. (2007). An introduction to the Rasch measurement model: An example using the Hospital Anxiety and Depression Scale (HADS). British Journal of Clinical Psychology, 46(1), 1–18. 10.1348/014466506X96931

Preiss-Farzanegan, S. J., Chapman, B., Wong, T. M., Wu, J., & Bazarian, J. J. (2009). The Relationship Between Gender and Postconcussion Symptoms After Sport-Related Mild Traumatic Brain Injury. PM&R, 1(3), 245–253. 10.1016/j.pmrj.2009.01.011

Randolph, C., Millis, S., Barr, W. B., McCrea, M., Guskiewicz, K. M., Hammeke, T. A., & Kelly, J. P. (2009a). Concussion Symptom Inventory: An Empirically Derived Scale for Monitoring Resolution of Symptoms Following Sport-Related Concussion. Archives of Clinical Neuropsychology, 24(3), 219–229. 10.1093/arclin/acp025

Rutherford, William H., Merrett, John D., & Mcdonali, John R. (1977). SEQUELÆ OF CONCUSSION CAUSED BY MINOR HEAD INJURIES. The Lancet, 309(8001), 1–4. 10.1016/S0140-6736(77)91649-X

Schatz, P., Moser, R. S., Covassin, T., & Karpf, R. (2011). Early Indicators of Enduring Symptoms in High School Athletes With Multiple Previous Concussions. Neurosurgery, 68(6), 1562–1567. 10.1227/NEU.0b013e31820e382e

Shaikh, N., Theadom, A., Siegert, R., Hardaker, N., King, D., & Hume, P. (2021). Rasch analysis of the Brain Injury Screening Tool (BIST) in mild traumatic brain injury. BMC Neurology, 21(1), 376. 10.1186/s12883-021-02410-6

Sinnott, A. M., Eagle, S. R., Kochick, V., Bricker, I. R., Collins, M. W., Sparto, P. J., Flanagan, S. D., Elbin, R. J., Connaboy, C., & Kontos, A. P. (2023). Test-Retest, Interrater Reliability, and Minimal Detectable Change of the Dynamic Exertion Test (EXiT) for Concussion. Sports Health: A Multidisciplinary Approach, 15(3), 410–421. 10.1177/19417381221093556

Stemper, B. D. (2022). Sport-related concussion: The role of repetitive head impact exposure. In Cellular, Molecular, Physiological, and Behavioral Aspects of Traumatic Brain Injury (pp. 29–40). Elsevier. 10.1016/B978-0-12-823036-7.00023-2

Tennant, A., & Conaghan, P. G. (2007). The Rasch measurement model in rheumatology: What is it and why use it? When should it be applied, and what should one look for in a Rasch paper? Arthritis Care & Research, 57(8), 1358–1362. 10.1002/art.23108

the EORTC Quality of Life Group and the Quality of Life Cross-Cultural Meta-Analysis Group, Scott, N. W., Fayers, P. M., Aaronson, N. K., Bottomley, A., De Graeff, A., Groenvold, M., Gundy, C., Koller, M., Petersen, M. A., & Sprangers, M. A. (2010a). Differential item functioning (DIF) analyses of health-related quality of life instruments using logistic regression. Health and Quality of Life Outcomes, 8(1), 81. 10.1186/1477-7525-8-81

Tutz, G., Schauberger, G., & Berger, M. (2018). Response Styles in the Partial Credit Model. Applied Psychological Measurement, 42(6), 407–427. 10.1177/0146621617748322

Wallace, J., Covassin, T., & Beidler, E. (2017a). Sex Differences in High School Athletes’ Knowledge of Sport-Related Concussion Symptoms and Reporting Behaviors. Journal of Athletic Training, 52(7), 682–688. 10.4085/1062-6050-52.3.06

Wilmoth, K., Magnus, B. E., McCrea, M. A., & Nelson, L. D. (2020). Preliminary Validation of an Abbreviated Acute Concussion Symptom Checklist Using Item Response Theory. The American Journal of Sports Medicine, 48(12), 3087–3093. 10.1177/0363546520953440

Yengo-Kahn, A. M., Hale, A. T., Zalneraitis, B. H., Zuckerman, S. L., Sills, A. K., & Solomon, G. S. (2016). The Sport Concussion Assessment Tool: A systematic review. Neurosurgical Focus, 40(4), E6. 10.3171/2016.1.FOCUS15611

Zuckerman, S. L., Apple, R. P., Odom, M. J., Lee, Y. M., Solomon, G. S., & Sills, A. K. (2014). Effect of sex on symptoms and return to baseline in sport-related concussion: Clinical article. Journal of Neurosurgery: Pediatrics, 13(1), 72–81. 10.3171/2013.9.PEDS13257

Zuckerman, S. L., Kerr, Z. Y., Yengo-Kahn, A., Wasserman, E., Covassin, T., & Solomon, G. S. (2015). Epidemiology of Sports-Related Concussion in NCAA Athletes From 2009-2010 to 2013-2014: Incidence, Recurrence, and Mechanisms. The American Journal of Sports Medicine, 43(11), 2654–2662. 10.1177/0363546515599634

